# Development of a Highly Sensitive Serum Neurofilament Light Chain Assay on an Automated Immunoassay Platform

**DOI:** 10.1101/2022.04.17.22273097

**Authors:** Stephen Lee, Tatiana Plavina, Carol M Singh, Kuangnan Xiong, Xiaolei Qiu, Richard A Rudick, Peter A Calabresi, Lauren Stevenson, Danielle Graham, Denitza Raitcheva, Christopher Green, Madeleine Matias, Arejas J Uzgiris

## Abstract

**Background:** Neurofilament light chain (NfL) is an axonal cytoskeletal protein that is released into the extracellular space following neuronal or axonal injury associated with neurological conditions such as multiple sclerosis (MS), amyotrophic lateral sclerosis (ALS), and other diseases. NfL is detectable in cerebrospinal fluid (CSF) and blood. Numerous studies in MS have demonstrated that NfL correlates with disease activity, predicts disease progression, and is reduced by treatment with MS disease-modifying drugs, making NfL an attractive candidate to supplement existing clinical and imaging measures in MS. However, for NfL to achieve its potential as a clinically useful biomarker for clinical decision-making or drug development, a standardized, practical, widely accessible assay is needed. Our objective was to validate the analytical performance of the novel serum neurofilament light (sNfl) assay on the ADVIA Centaur® XP immunoassay system.

**Methods:** The research assay was evaluated on the ADVIA Centaur XP immunoassay system from Siemens Healthineers. The lower limit of quantitation (LLoQ), intra-assay variation, assay range, cross-reactivity with neurofilament medium and heavy chains, and effect of interfering substances were determined. NfL assay values in serum and CSF were compared with radiological and clinical disease activity measures in patients with MS and ALS, respectively. This assay was further optimized to utilize serum, plasma, and CSF sample types and transferred to Siemens’ CLIA laboratory, where it was analytically validated as a laboratory-developed test.

**Results:** In this study, a LLoQ of 1.85 pg/mL, intra-assay variation of <6%, and an assay range of up to 646 pg/mL were demonstrated. A cross-reactivity of <0.7% with neurofilament medium and heavy chains was observed, and the assay was not significantly affected by various interfering substances encountered in clinical specimens. Serum and CSF NfL assay values were associated with radiological and clinical disease activity measures in patients with MS and ALS, respectively.

**Conclusion:** The analytical performance of the NfL assay fulfilled all acceptance criteria; therefore, we believe the assay is acceptable for use in both research and clinical practice settings to determine elevated sNfL levels in patients.

## INTRODUCTION

One challenge for clinicians in managing neurodegenerative diseases is the lack of biomarkers that provide a quantitative measure of underlying disease severity and activity and prove useful to monitor effectiveness of disease-modifying therapies (DMTs) (1-4). Molecular biomarkers that originate in the central nervous system (CNS), which is shielded by the blood-brain barrier, have been previously thought to be inaccessible to blood-based testing. With recent advances in diagnostic technology, measuring very low levels of such biomarkers is now possible using routine clinical laboratory platforms.

Neurofilament light chain (NfL) is a scaffolding protein found specifically in the neuronal cytoskeleton and is released into the extracellular space following axonal degeneration (5-7). As such, it is a promising biomarker that may have applications for stratifying disease severity, monitoring activity or progression of neurodegenerative disorders, and determining efficacy of treatments (8).

NfL levels are known to correlate with the extent of axonal damage in a variety of neurological disorders (9). For multiple sclerosis (MS), it has been reported that baseline serum NfL (sNfL) is a predictor of long-term brain atrophy, development of new T2 lesions, T2 lesion volume, gadolinium (Gd+) lesions, and increased likelihood of progression from radiologically isolated syndromes or clinically isolated syndromes to clinically definite MS (10, 11). In addition, sNfL levels are higher and more variable in patients with evidence of active MS and decrease with a DMT (11). Multiple reports have shown that sNfL levels are responsive to treatment with MS DMTs (12-15). Similarly, NfL levels from the cerebrospinal fluid (CSF) of patients with amyotrophic lateral sclerosis (ALS) predict disease severity before it is clinically manifest (16).

It is thought that the incorporation of NfL measurements into clinical decision-making may improve patient outcomes by allowing detection of aggressive disease earlier, and by providing more effective monitoring to inform choice of appropriate therapeutic regimen and other care measures. Incorporation of NfL measurements into drug development may allow informative enrollment into clinical trials and a sensitive measurement of treatment effect, thereby reducing required sample sizes for early stage trials.

To incorporate NfL testing in clinical practice, measurement of NfL levels will need to be standardized and accessible. Most other assays used to generate evidence for the utility of NfL have run on research use only platforms (17). The goal of this report is to describe development and validation of a novel NfL assay that runs on the ADVIA Centaur immunoassay system, a routine clinical laboratory platform.

## MATERIALS AND METHODS

### Assay Development Overview

A summary of the key development and optimization studies for the NfL assay is shown in **Table 1**. In short, multiple antibody candidates were screened, and one antibody pair was selected for assay development. Antibodies were conjugated to biotin and an acridinium ester (tracer) for compatibility with Siemens immunoassay analyzers. Optimal assay formulations, critical assay parameters, and control systems were established prior to assessing analytical performance.

**Table 1.** Summary of NfL assay development and optimization activities. CLIA, Clinical Laboratory Improvement Amendments; NfL, neurofilament light chain.

|  |
| --- |
| 1. Screening and selection of capture and detection antibodies |
| 2. Prototyping with capture and detection antibodies in reagents on instrument |
| 3. Preliminary feasibility studies |
| 4. Optimization of assay parameters |
| 5. Control system development |
| 6. Further development of additional sample types and technology transfer to CLIA laboratory |

### NfL Research Use Assay

The design of this NfL Assay is immunometric — using solid-phase magnet bead capture with one antibody, and direct acridinium ester chemiluminometric detection with the other. An initial NfL assay prototype was implemented and evaluated as a research assay on the ADVIA Centaur immunoassay system (**Figure 1**). Using the same materials, a further universal sample type (plasma/serum/CSF) research use NfL assay was optimized for, and analytically validated on, the Atellica Solution immunoassay system. The light reagent comprises a different monoclonal anti-NfL antibody labeled with a proprietary acridinium ester (AE) for chemiluminescent detection. Accumulated light signal is related to NfL concentration in the sample. Calibrators and control materials were also developed to enable reliable, highly sensitive, and quantitative reporting. Calibrators across the range of the assay were used to generate a standard curve for the research assay.

**Figure 1.**
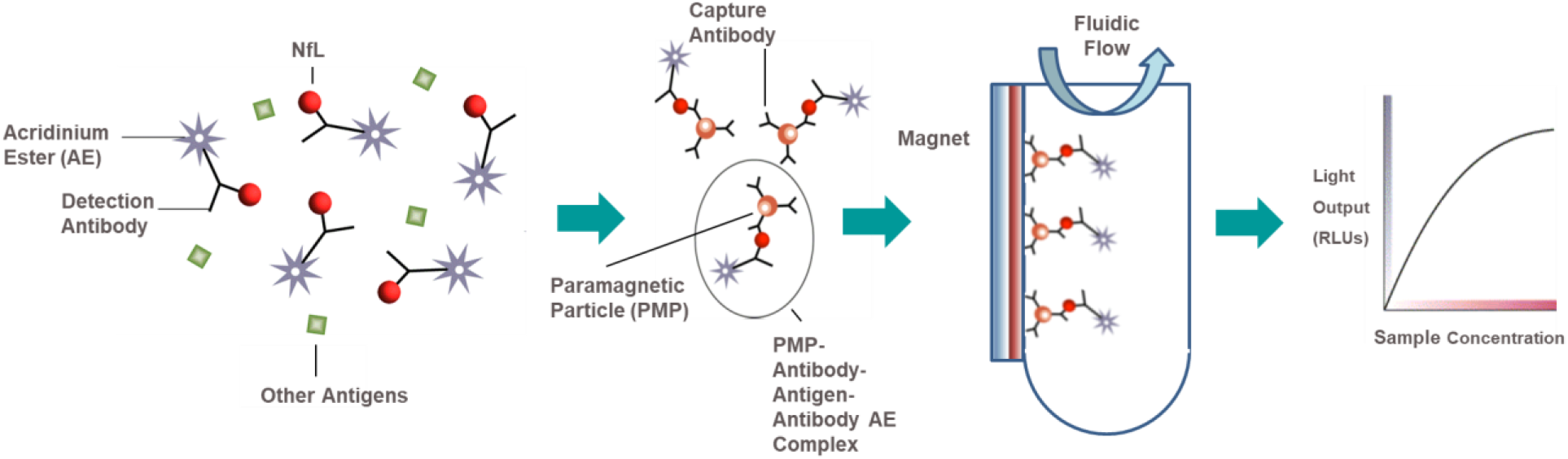
Acridinium ester-based automated immunoassay workflow. As implemented on the Siemens Centaur^®^ and Atellica^®^ testing platform. AE, acridinium ester; PMP, paramagnetic particle; RLUs, relative luminescence units.

### Analytical Samples and Other Materials

Off-the-clot human serum pools (Access Biologicals, Vista, CA), K2 EDTA human plasma pools (Access Biologicals, Vista, CA), individual and matched serum (Access Biologicals, Vista, CA and BioIVT, Shirley, NY), K2 EDTA plasma (BioIVT, Shirley, NY), and lithium heparin plasma (BioIVT, Shirley, NY) were sourced. CSF samples were acquired from commercial sources (BioIVT, Shirley, NY). To establish the detection capabilities of the NfL assay at the low end of the assay range, contrived samples were utilized and prepared by using NfL-depleted serum, plasma, or CSF. NfL-depleted matrixes were prepared by immuno-absorption. To prepare samples with higher NfL concentrations, recombinant human NfL and endogenous NfL from CSF were used as spikers.

Two tri-level serum-based quality control (QC) sets known herein as Siemens NfL QCs were prepared using off-the-clot human serum. One set known as endogenous QCs (EQC 1-3) consisted of one neat (or unspiked) serum pool and two other serum pools spiked with CSF to target NfL concentrations of 16 and 50 pg/mL. A second set known as recombinant QCs (RQC1-3) was prepared by spiking serum with recombinant human NfL to target NfL concentrations of 16, 50, and 450 pg/mL. The CSF sample used to spike endogenous QC materials originated from a donor with ALS. A plasma tri-level set for precision studies with the laboratory-developed test (LDT) version of the assay consisted of one neat (or unspiked) K2 EDTA plasma pool and two other plasma pools spiked to target NfL concentrations of 50 and 400 pg/mL with recombinant human NfL. Serum, plasma, and CSF samples sourced from individual donors were used for parallelism experiments. Parallelism samples were diluted serially with NfL depleted matrices to lower concentration levels.

### Assay Precision

Repeatability and within-lab precision were assessed according to Clinical and Laboratory Standards Institute (CLSI) Document EP05-A3 (18) using a 20-day x 2 run x 2 replicate design with one reagent lot tested on one instrument. Aliquots of the six Siemens NfL QCs were prepared and frozen at −70 °C prior to the start of the study. On the morning of each testing day, an aliquot was thawed to room temperature, mixed by inversion, then transferred to a sample rack for duplicate testing. This process was repeated in a second run (at least 2 hours after the first run) on the same testing day using a fresh aliquot. In total, each serum sample generated 80 measurements over 40 independent runs.

### Interfering Substances

Potential interferents such as Intralipid (Sigma-Aldrich, St. Louis, MO), Cholesterol (Lee Biosolutions, Maryland Heights, MO), Human Serum Albumin (Lee Biosolutions, Maryland Heights, MO), Human Hemoglobin (Lee Biosolutions, Maryland Heights, MO), Indirect Bilirubin (Conjugate; Lee Biosolutions, Maryland Heights, MO), Direct Bilirubin (Lee Biosolutions, Maryland Heights, MO), Rheumatoid Factor Serum (Lee Biosolutions, Maryland Heights, MO), and Biotin (Sigma-Aldrich, St. Louis, MO) were spiked at targeted to minimum concentrations recommended by CLSI EP37 in three of the Siemens NfL QCs that spanned low, medium, and high levels of NfL.(19) Control samples that did not contain interferent were prepared by spiking the same samples with the storage buffer of each interferent. Interference was expressed as absolute percent bias between the mean test and control sample results.

### Analytical Specificity

Specificity was determined by spiking two other neurofilaments, neurofilament heavy chain (NfH) and neurofilament medium chain (NfM), into three Siemens NfL QCs and NfL-depleted serum. Purified bovine NfM and NfH (Origene, Rockville, MD) were each spiked into four samples spanning the NfL assay range (0–500 pg/mL) at target concentrations of 1000 pg/mL from stocks 100-fold more concentrated. Control samples were prepared by spiking the same four NfL samples with the storage buffer used to reconstitute NfM and NfH. Cross-reactivity was calculated as the percent difference between the mean test and control sample results, with respect to the test analyte concentration.

### Analytical Sensitivity

Detection capability for limit of blank (LoB), limit of detection (LoD), and limit of quantitation was estimated in accordance with CLSI Document EP17-A2 and CLSI Document EP05-A3 (18, 20). Four different NfL-depleted human serum pools served as LoB samples and four NfL-depleted serum pools each spiked with neat pooled human serum at target concentrations of 1–4 times the LoB served as LoD samples. A human serum pool was diluted with NfL depleted serum down to the LoB to yield 6 additional LLoQ panel samples. A human serum pool spiked with endogenous NfL from CSF to a target concentration of 16 pg/mL served as the highest LoQ sample. All LoB and LoD samples were assayed in replicates of five per run daily over 3 days with two reagent lots (60 total blank and 60 total low-level sample measurements per lot) on one instrument. LoB was calculated as the 95th percentile for the ranked results. LoD was determined parametrically from the low-level samples only using the equation: LoD = LoB + CpSDL, where Cp is a multiplier to give 95th percentile of a normal distribution. LLoQ samples (target concentrations from assay LoB to 15-fold above the LoB) were tested two runs a day, four replicates per run for 5 days with two different reagent lots (40 total measurements per sample per lot) on one instrument (**Figure 2**). LLoQ was determined using the precision profile method, where the LLoQ is calculated at the concentration corresponding to within-lab coefficient of variation of 20%.

**Figure 2.**
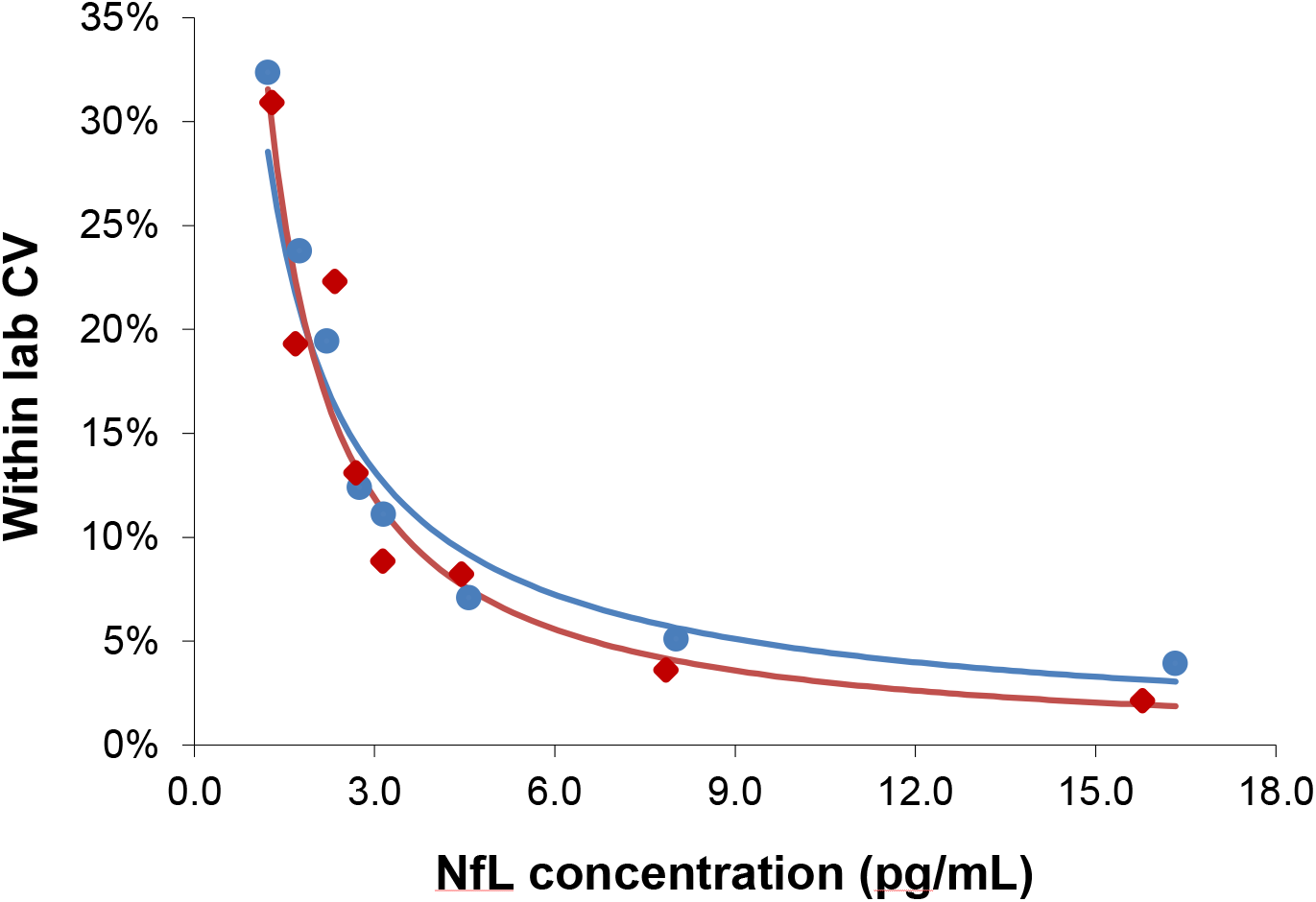
Lower limit of quantitation. Analytical samples at eight concentrations spanning the assay LoB (0.89 pg/mL) to 15-fold above the LoB. Samples generated from donor serum with endogenous NfL diluted in pooled donor serum first depleted of NfL using antibody immunoabsorption. Inter-run, inter-day, and inter-lot variation were tested (four replicates per sample; two runs per day over 5 days; two reagent lots). The lower limit of quantitation for Lots 1 and 2 were 1.84 and 1.85 pg/mL, respectively, at a precision cut-off of 20% CV. The blue and red regression lines correspond to Lots 1 and 2, respectively. CV, coefficient of variation; LoB, limit of blank; NfL, neurofilament light chain.

### Linearity

Assay linearity was tested according to CLSI Document EP06-A (21) with three replicates of nine samples across the 0–500 pg/mL range. The nine serum samples of evenly spaced NfL concentration were prepared by mixing two contrived serum pools at opposite ends of the NfL assay range (**Figure 3A**). The highest contrived serum pool was spiked with recombinant NfL to slightly above the assay measurement range. The low concentration pool was NfL-depleted serum pool. Linear regression analysis was performed between measured concentration and coded pool number.

**Figure 3.**
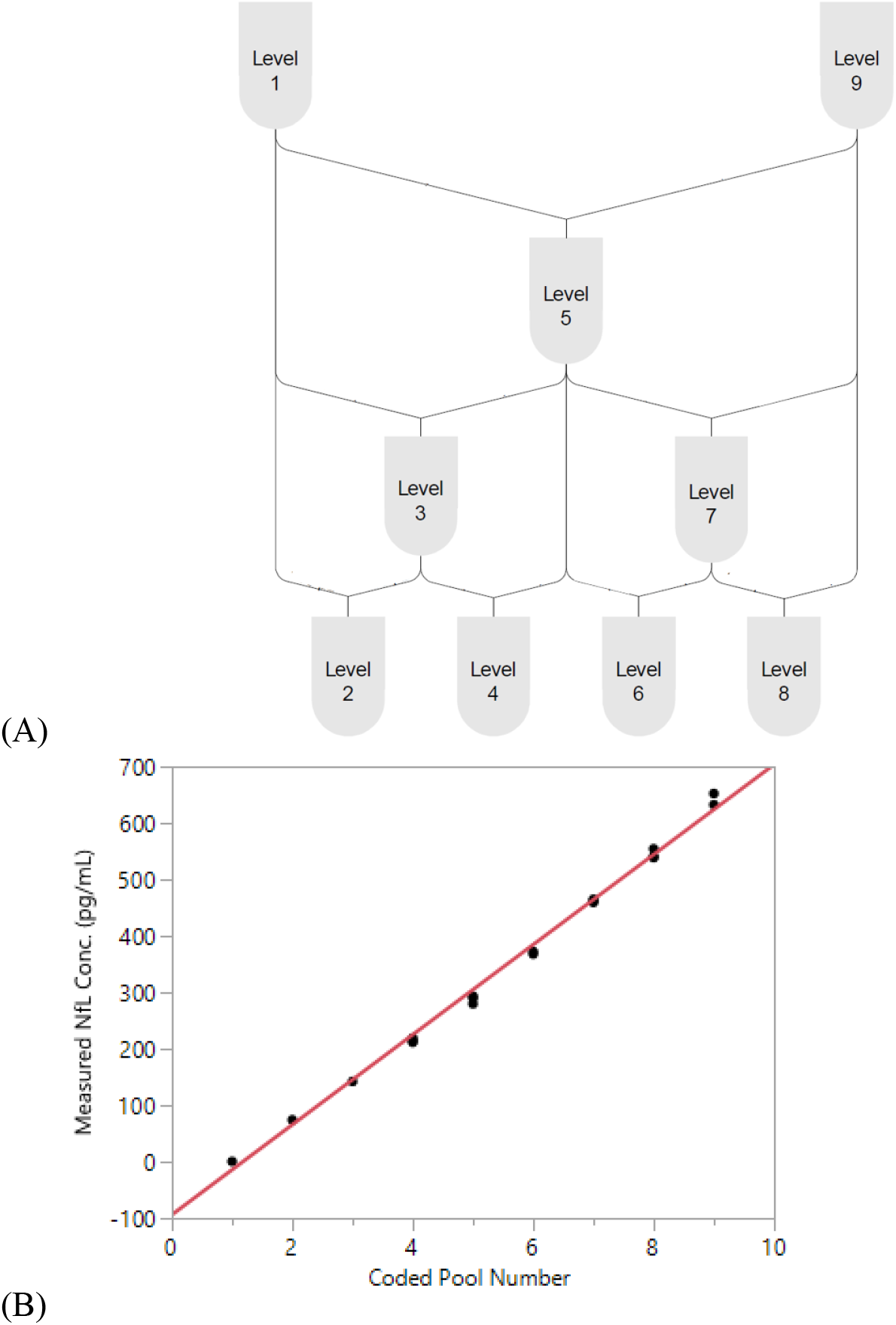
Serum linearity. (A) Sample stock pools with high (serum spiked with recombinant NfL to above ULoQ) and low (NfL-depleted serum below LLoQ) levels of NfL were used to prepare nine concentration levels evenly spaced across the assay range. (B) A regression between measured concentration and coded pool number was performed. LLoQ, lower limit of quantitation; NfL, neurofilament light chain; ULoQ, upper limit of quantitation.

### Parallelism

Parallelism was defined as a condition in which dilution of test samples does not result in biased measurements of the analyte concentration (with limits of 80–120% recovery). Parallelism was assessed using 10 serum samples from healthy individuals with relatively high NfL concentrations (range from 16– 35 pg/mL). Each sample was diluted 1:2, 1:4, and 1:8. Measured concentrations of the diluted samples were multiplied by their dilution factor and compared to their neat concentration by percentage of recovery.

### Spike Recovery

The spike recovery of recombinant NfL in serum was performed with two serum cohorts. The first cohort consisted of five individual samples, where one outlier was found. Therefore, a larger cohort of 50 individual donors was tested to determine the interference frequency. To assess percent recovery, each sample was spiked with recombinant NfL, and individual and mean percentage of recovery and were calculated and reported.

### Hook Effect

Hook (or prozone) effect was evaluated using a series of dilutions of a high NfL sample with two reagent lots. Recombinant NfL was spiked in serum to a target concentration of 500 ng/mL (1000 times the upper limit of quantification [ULoQ]) as a high sample. The high sample and a series of dilutions of the high sample were tested with a control sample targeted at the ULoQ in replicates of three. A hook effect was defined as any instance when the three times the standard deviation interval of the dilution overlaps the response of the control sample.

### Sample Stability and Handling

Stability of serum samples was assessed at both ambient temperature and after freeze-thaws. Four individual donor serum samples were tested after storage on the bench at room temperature (20–25 °C) for 4, 8, 24, and 48 hours. Additionally, separate aliquots from the same four individual donors were also assessed for stability up to five freeze/thaw cycles (**Supplementary Figure 1**).

### Reagent Storage Stability

Siemens immunoassay reagents were manufactured in consumable packs compatible with its automated analyzers. The stability of reagents stored at their recommended storage conditions of 2-8°C was assessed by monitoring the measured drift of QC samples. QCs were tested on Day 0, 3, 7, 10, 14, 17, 21, 24, 28, 31, 72, 123. Linear regression analysis of concentration versus time for each sample was performed. If the regression line slope was not statistically significant (*P*-value > 0.05), the stability duration estimate for that sample was taken as the last time point tested. If the slope was significant, the stability duration estimate was set as the time at which a one-sided 95% confidence interval about the regression line intersects with the allowable drift acceptance criterion for that sample. Allowable drift acceptance criterion was 20% for QC samples.

### Reagent Onboard Stability

The in-use stability of a reagent pack stored onboard was assessed and a calibration interval was established. Three NfL samples (one endogenous with low NfL concentration; two recombinant with NfL medium and high concentrations) were tested at 0-, 3-, 7-, 10-, 14-, 17-, 21-, 24-, 28-, and 31-day time points on both a fresh pack (stored at 2–8 °C) and in-use pack (stored onboard). Linear regression analysis of concentration versus time for each sample was performed similarly to reagent pack shelf-life stability. Allowable drift acceptance criterion was 20% for QC samples.

### Method Comparison

Method comparison studies were performed in accordance with CLSI Document EP09-A3. Correlation of NfL results with the SIMOA NfL-light^®^ Advantage Kit (Quanterix, Billerica, MA) was examined using 458 clinical serum samples from the MSPATHS biorepository (*n* = 241) (22) and the ADVANCE clinical trial (*n* = 217) (11). Each sample was diluted three-fold with Siemens NfL Sample Diluent and assayed in singlicate on one instrument. NfL values below the LLoQ were excluded in the method comparison analysis.

### Clinical Application

Serum samples and MRI images were collected from patients with MS enrolled in the ADVANCE study, a randomized, multicenter, double-blind, placebo-controlled study assessing the efficacy and safety of peginterferon beta-1a for patients with relapsing-remitting MS (11). NfL levels were measured using the assay described herein, and the number of new T2 lesions was derived from MRI images. CSF samples from healthy controls and patients with a diagnosis of clinically definite ALS were obtained from a research agreement between Biogen, Inc. (Cambridge, MA) and Iron Horse Diagnostics (Phoenix, AZ).

### Additional Studies (Only Performed for the Laboratory Developed Test)

#### Specimen Equivalence

Specimen equivalence was assessed for three types of collection tubes: serum, K2 EDTA plasma, and lithium heparin plasma. Matched tube types from 40 individuals were tested without dilution.

### Assay Precision (with K2EDTA Plasma)

Repeatability and within-lab precision were assessed using a 5-day x two run x two replicate design with two reagent lots tested on one instrument. In short, aliquots of the three plasma samples spanning the assay range were prepared and frozen at –70°C prior to the start of the study. One sample was pooled human plasma, and two other samples were created from the same plasma pool spiked with recombinant NfL to target levels of 50 and 400 pg/mL. On the morning of each testing day, an aliquot was thawed to room temperature, mixed by inversion, then transferred to a sample rack for duplicate testing. This process was repeated in a second run (at least 2 hours after the first run) of the same testing day using a fresh aliquot. In total, each plasma sample had 20 measurements over 10 independent runs.

### Parallelism (K2EDTA and Li Heparin Plasma)

Parallelism was assessed for plasma tube types using 10 samples: five K2EDTA and five lithium heparin samples from healthy individuals with relatively high normal NfL concentrations. Each sample was diluted 1:2, 1:4, 1:8, and 1:10. Measured concentrations of the diluted samples were multiplied by their dilution factor and compared to their neat concentration by percent recovery. Parallelism was demonstrated if recovery was within 80–120%.

### Parallelism (CSF)

Seven individual CSF samples were serially diluted 10-, 20-, 40-, 80-, 160-, and 400-fold using NfL Sample Diluent. CSF was diluted with serum at least 10-fold to ensure the test-matrix was primarily serum-based and compatible with the NfL assay. Relative recovery was calculated in comparison to dilution-corrected concentration tested at 10-fold.

## RESULTS

### Analytical Performance (For Research Assay with Serum Only)

#### Precision

Repeatability and within-lab precision for Siemens Healthineers NfL QCs are summarized in **Table 2**. The within-lab percent coefficient of variation was less than 6% during the 20-day period.

**Table 2.** Precision with NfL research assay (serum). CV, coefficient of variation, NfL, neurofilament light chain, SD, standard deviation.

|  |  | Repeatability |  | Within-Lab Precision |  |
| --- | --- | --- | --- | --- | --- |
| Sample | Mean (pg/mL) | SD (pg/mL) | CV | SD (pg/mL) | CV |
| Endogenous Level 1 (EQC1) | 7.65 | 0.26 | 3.4% | 0.39 | 5.1% |
| Endogenous Level 2 (EQC2) | 17.23 | 0.57 | 3.3% | 0.76 | 4.4% |
| Endogenous Level 3 (EQC3) | 54.18 | 0.96 | 1.8% | 1.66 | 3.1% |
| Recombinant Level 1 (RQC1) | 17.89 | 0.39 | 2.2% | 0.58 | 3.2% |
| Recombinant Level 2 (RQC2) | 54.95 | 0.84 | 1.5% | 1.37 | 2.5% |
| Recombinant Level 3 (RQC3) | 438.03 | 7.49 | 1.7% | 12.17 | 2.8% |

#### Interference

Interferent concentration tested and absolute percent bias for three Siemens NfL QCs are summarized in **Table 3**. Significant interference was considered absolute bias ≥10% for all NfL levels. No significant interference was observed for the interferents except hemoglobin at 500 mg/dL (bias not shown). Lower concentrations of hemoglobin at 200 mg/dL showed no significant interference.

**Table 3.**
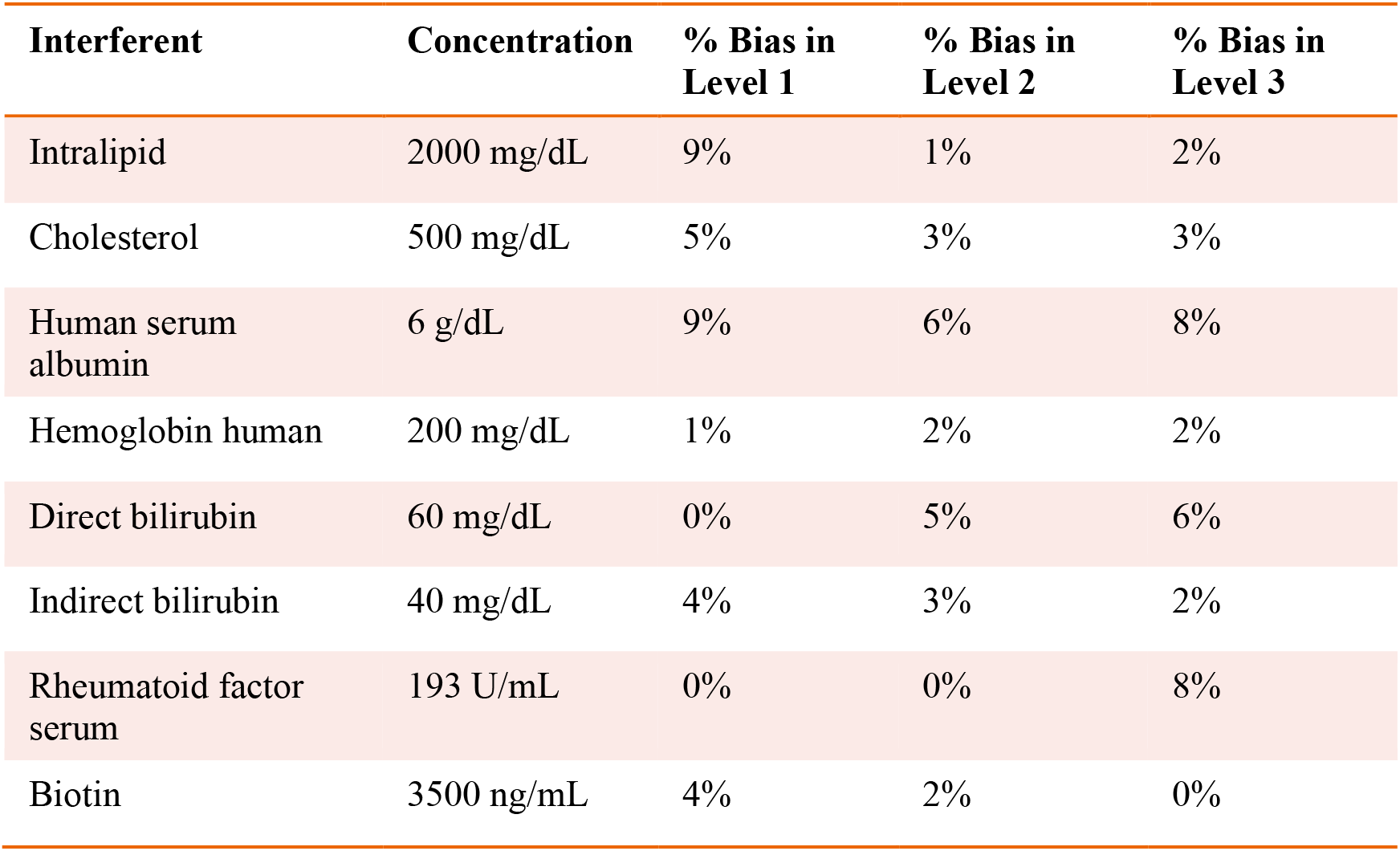
Interference testing. Siemens NfL assay performance is not affected by following interferents. NfL, neurofilament light chain.

| Interferent | Concentration | % Bias in Level 1 | % Bias in Level 2 | % Bias in Level 3 |
| --- | --- | --- | --- | --- |
| Intralipid | 2000 mg/dL | 9% | 1% | 2% |
| Cholesterol | 500 mg/dL | 5% | 3% | 3% |
| Human serum albumin | 6 g/dL | 9% | 6% | 8% |
| Hemoglobin human | 200 mg/dL | 1% | 2% | 2% |
| Direct bilirubin | 60 mg/dL | 0% | 5% | 6% |
| Indirect bilirubin | 40 mg/dL | 4% | 3% | 2% |
| Rheumatoid factor serum | 193 U/mL | 0% | 0% | 8% |
| Biotin | 3500 ng/mL | 4% | 2% | 0% |

#### Specificity

Cross-reactivity of the Siemens NfL assay with purified NfM and NfH was below 0.7% for four different serum samples with NfL concentrations spanning the assay range (**Table 4**).

**Table 4.** Cross-reactivity assessment. No cross-reactivity observed with NfM or NfH. ND reported when the concentration difference between test and control samples is below the LoD. LoD, limit of detection; ND, not detectable; NfH, neurofilament heavy chain; NfM, neurofilament medium chain.

| Test substance | Concentration (pg/mL) | % Cross-reactivity |  |  |  |
| --- | --- | --- | --- | --- | --- |
|  |  | Target 0 pg/mL NfL-depleted serum | Target 7.65 pg/mL (Endogenous Level 1) | Target 17.23 pg/mL (Endogenous Level 2) | Target 438.03 pg/mL (Recombinant Level 3) |
| NfM | 1000 | ND | 0.0146% | 0.0380% | 0.3705% |
| NfH | 1000 | ND | 0.0233% | 0.0011% | 0.6780% |

#### Sensitivity

The highest LoB, LoD, and LLoQ results among the two reagent lots are reported for the assay. Three out of four LoD samples (45 of 60 measurements) were used to determine the LoD. One LoD sample after completion of the study was excluded because the analyte concentration was too close to the LoB and could not be used for SD calculation. LoB was determined to be 0.89 pg/mL, and LoD was calculated as 1.49 pg/mL. LLoQ was determined using the precision profile method and equation of the power trendline fit and determined to be 1.85 pg/mL.

#### Linearity

Linearity of the NfL assay was observed across the range of 1–646 pg/mL. Linear regression results were R^2^ = 0.996 with *P*-value < 0.001 (**Figure 3B**).

#### Parallelism (Serum)

Parallelism was demonstrated with the research assay first in 10 individual sera with endogenous NfL levels ranging from 16 – 35 pg/mL. All dilutions recovered within 80-120% of the neat measurement of each sample after adjusting for dilution factor (**Figure 4**).

**Figure 4.**
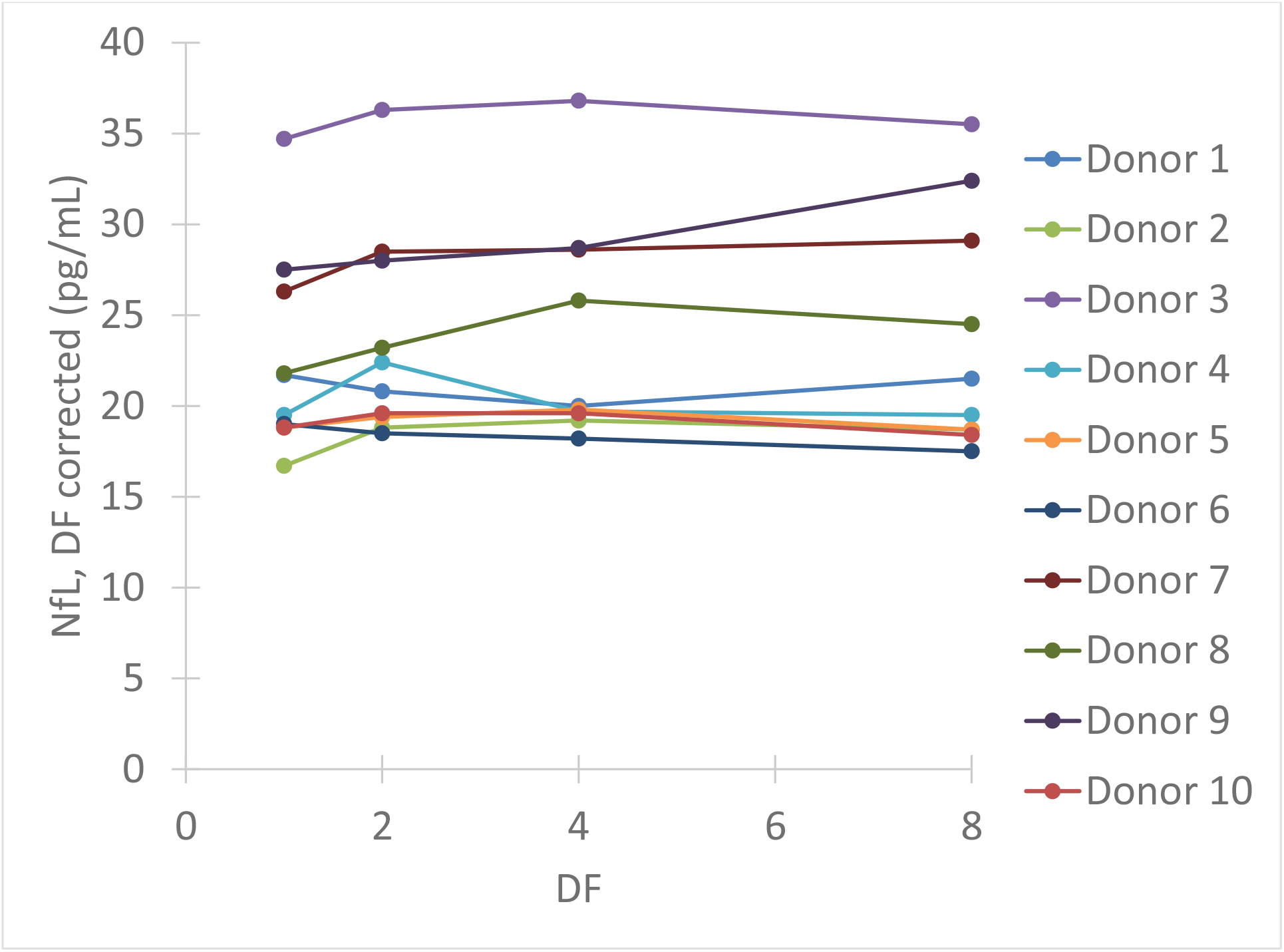
Serum parallelism. Ten individual serum samples with endogenous NfL levels >16 pg/mL (up to 35 pg/mL) were tested neat, and serially diluted 2X, 4X, and 8X using NfL sample diluent. DF, dilution factor; NfL, neurofilament light chain.

#### Spike Recovery

Spike recovery was within our acceptance criteria of 80–120% for more than 95% of the samples (53 out of 55) of the samples (not shown).

#### Hook Effect

The hook or prozone effect is a phenomenon where the formation of antibody-antigen immune complexes can be impaired when concentrations of the measurand (antigen or antibody depending on the type of assay) are very high. When there is a hook effect, there is a concentration point when the immunoassay measures less measurand when the measurand concentration is increasing, producing a hook shape on a graph of measurements. No hook effect was observed below 481 ng/mL for the two reagent lots tested (**Supplementary Figure 2**).

#### Sample Stability

Serum NfL stability was assessed at room temperature for up to 48 hours and over five freeze-thaw cycles. All samples were stable in these conditions as demonstrated by <5% difference from the control condition.

#### Reagent Pack Shelf-Life Stability

Assay reagents were found to be stable for up to 1 year (last time point tested) (not shown) when stored at recommended conditions of 2–8 °C. The shelf-life of the reagents continues to be monitored.

#### In-Use Stability

For all NfL samples tested across 31 days, no significant drift was observed (not shown, *P*-value > 0.05), demonstrating that reagent packs stored onboard are stable up to 28 days.

#### Method Comparison With SIMOA Assay from Quanterix

Analysis of MS patient serum samples (*n* = 418 above LLoQ) demonstrated high correlation (R^2^ = 0.907) between NfL results from the Siemens ADVIA Centaur XP and the Quanterix SIMOA platform (Figure 5).

**Figure 5.**
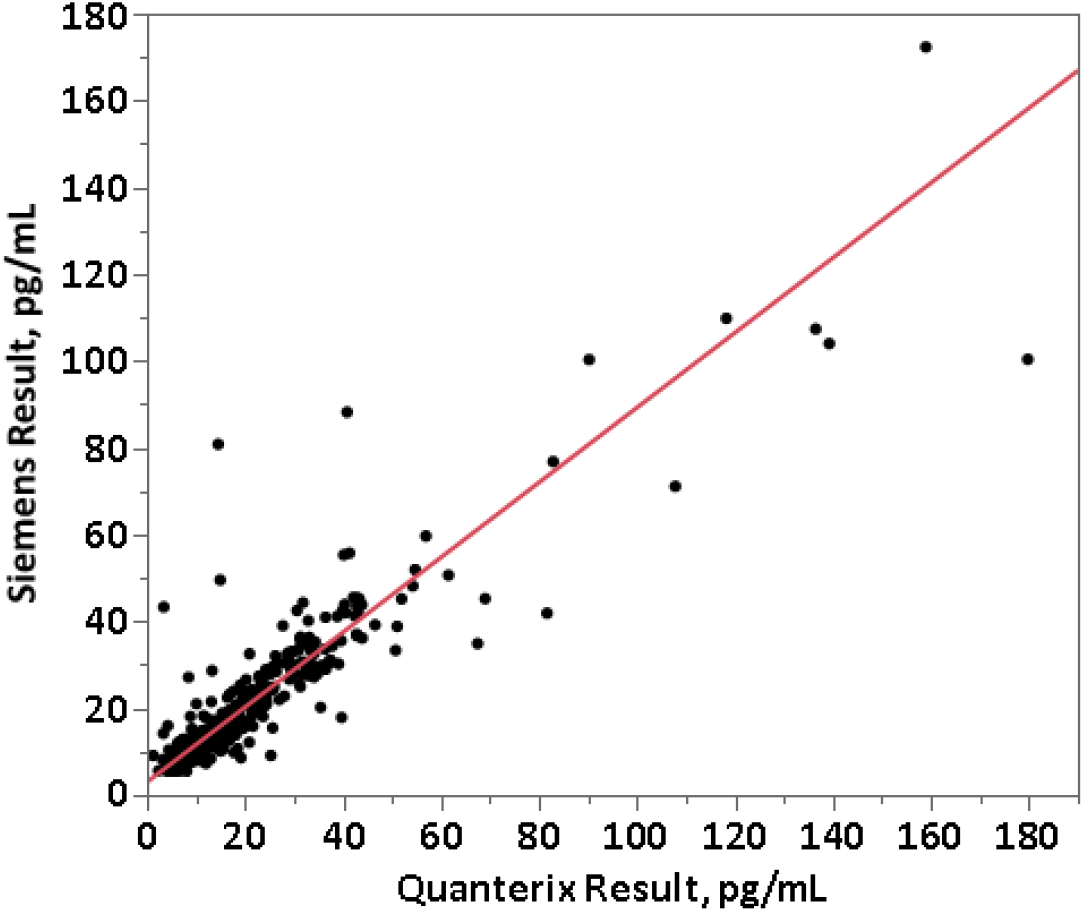
Method comparison between Siemens assay and Quanterix SIMOA assay. Data are from the MS PATHS and ADVANCE studies, selected over the range of Quanterix sNfL results and sample availability. sNfL, serum neurofilament light.

### Analytical Performance (for Laboratory Developed Test with Multiple Sample Types)

#### Specimen Equivalence

All three tube types demonstrated specimen equivalence (**Figure 6**). Linear fits for all tube type combination comparisons were within acceptance criteria of a slope equal to 1.0 ± 0.1 and y-intercept less than or equal to the LLoQ of the NfL assay (*P*-values < 0.0001).

**Figure 6.**
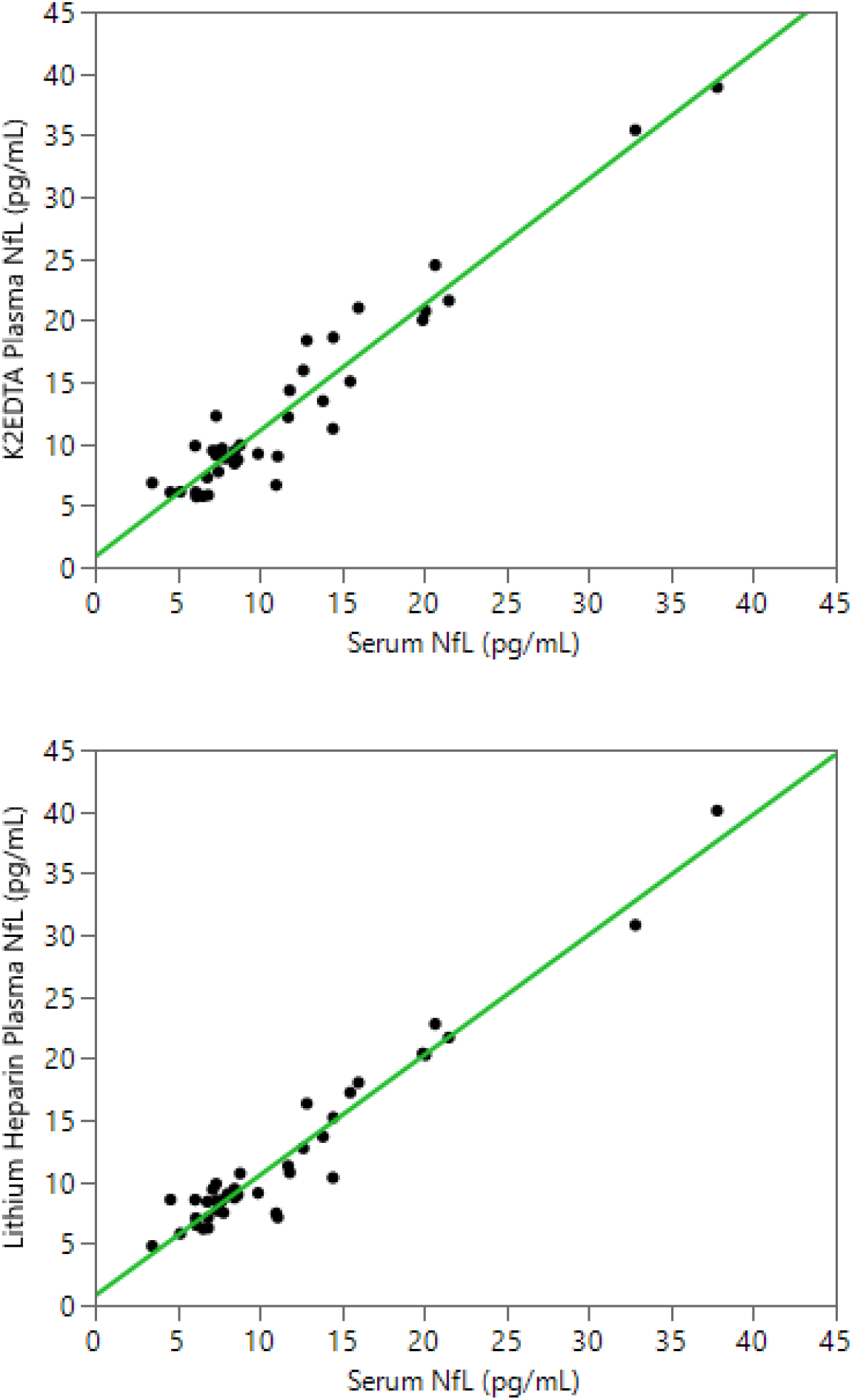
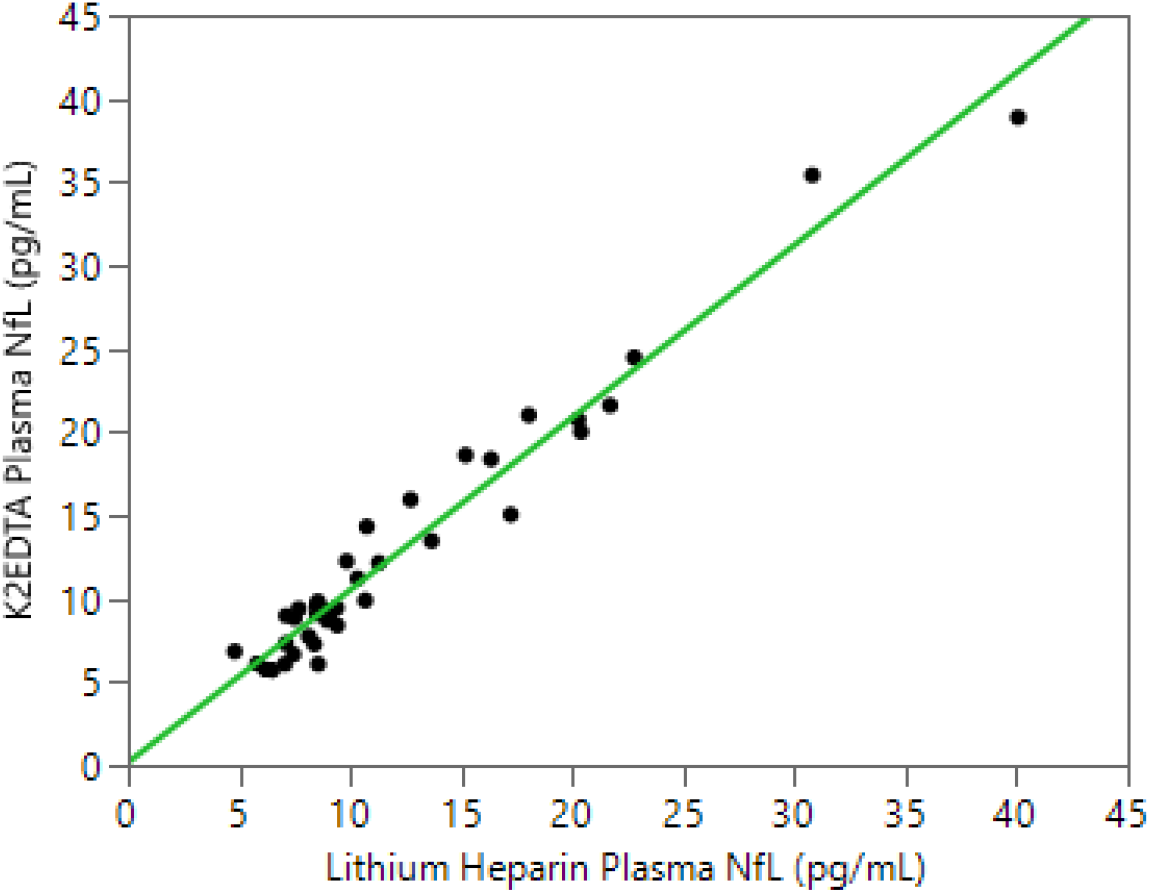
Equivalence of serum and plasma NfL levels in the LDT implementation of the NfL assay. LDT, laboratory-developed test; NfL, neurofilament light chain.

#### Precision with K2 EDTA Plasma

Repeatability and within-lab precision for the three tested plasma samples are summarized in **Table 5**. Within-lab percent coefficient of variation was ≤5.8% over the 5-day period for both reagent lots.

**Table 5.** Precision with LDT version of NfL assay (K2 EDTA plasma). CV, coefficient of variation, LDT, laboratory-developed test; NfL, neurofilament light chain; SD, standard deviation.

|  |  |  | Repeatability |  | Within-lab precision |  |
| --- | --- | --- | --- | --- | --- | --- |
| Sample | Reagent lot | Mean (pg/mL) | SD (pg/mL) | CV | SD (pg/mL) | CV |
| Plasma Level 1 | Lot 1 | 10.1 | 0.3 | 2.8 | 0.4 | 3.9 |
| Plasma Level 2 |  | 45.7 | 2.1 | 4.7 | 2.4 | 5.3 |
| Plasma Level 3 |  | 346.3 | 16.4 | 4.7 | 19.8 | 5.7 |
| Plasma Level 1 | Lot 2 | 9.9 | 0.2 | 2.4 | 0.4 | 4.2 |
| Plasma Level 2 |  | 46.0 | 2.4 | 5.2 | 2.7 | 5.8 |
| Plasma Level 3 |  | 325.0 | 14.0 | 4.3 | 17.0 | 5.2 |

**Table 6.** Summary of analytical validation of LDT version of NfL assay. CLSI, Clinical and Laboratory Standards Institute; CSF, cerebrospinal fluid; MS, multiple sclerosis; NfL, neurofilament light chain.

| Characteristic | Serum | Plasma | CSF |
| --- | --- | --- | --- |
| Reportable range | 3.9–500 pg/mL | 4.9–477 pg/mL (*K2 EDTA*)<br>2.4–549 pg/mL (*Lithium Heparin*) | 85.5–25,700 pg/mL |
| Reproducibility | 4.9–8.4% | 7.7–18.1% (*K2 EDTA*)<br>3.5–16.3% (*Lithium Heparin*) | 4.0–16.5% |
| Method comparison | *Quanterix vs Atellica® platform:*<br>Avg Quantitation Difference: –8%;<br>Pearson correlation: R=0.995<br>*ADVIA Centaur XP vs. Atellica® platform:*<br>Average Quantitation Difference: 9%;<br>Pearson correlation: R=1.0 | Tested for serum only; specimen equivalence was established based on precision and accuracy (CLSI EP35) | *Quanterix vs. Atellica® platform:*<br>Avg Quantitation Difference: –29%;<br>Pearson correlation: R=0.994<br>*ADVIA Centaur XP vs Atellica® platform:*<br>Average Quantitation Difference: -13.2%;<br>Pearson correlation: R=0.996 |
| Specimen handling | NfL is stable under the following conditions:<br>Up to 6 freeze/thaw cycles<br>Up to 1 week at room temperature<br>Up to 2 weeks refrigerated<br>Frozen at –20 °C for 1 year<br>Frozen at –80 °C for 1 year | NfL is stable under the following conditions:<br>Up to 6 freeze/thaw cycles<br>Up to 1 week at room temperature<br>Up to 2 weeks refrigerated<br>Frozen at –20 °C: 3 months (K2 EDTA) and 6 months (Lith Hep)<br>Frozen at –80 °C: 6 months (K2 EDTA) and 1 year (Lith Hep) | NfL is stable under the following conditions:<br>Up to 6 freeze/thaw cycles<br>Up to 1 week at room temperature<br>Up to 1 week refrigerated<br>Frozen at –20 °C for 3 months<br>Frozen at –80 °C for 1 year |
| Drug interference | Drug interference testing was performed using the following drugs used to treat patients with Alzheimer's and MS:<br>Donepezil<br>Rivastigmine<br>Memantine<br>Galantamine<br>Citalopram<br>Mirtazapine<br>Sertraline<br>Bupropion<br>Duloxetine<br>Imipramine<br>Ibuprofen<br>Siponimod<br>Acetaminophen<br>Aspirin<br>Beta interferon 1a<br>Beta interferon 1b<br>Fingolimod<br>Dimethyl fumarate<br>Teriflunomide<br>Ocrelizumab<br>Mitoxantrone<br>Caldribine<br>Alemtuzumab<br>Glucose<br>Drug interference (±20%) was observed in the presence of Mitoxantrone at concentrations greater than 0.113 mg/dL. | Tested for serum only; specimen equivalence was established based on precision and accuracy (CLSI EP35) | Tested for serum only; specimen equivalence was established based on precision and accuracy (CLSI EP35) |

#### Plasma Parallelism

Parallelism was demonstrated in matched K2 EDTA and lithium heparin plasma collected samples from 5 individuals (**Figure 7**). Endogenous levels ranged from 15.1 to 38.4 pg/mL and from 16.7 to 38.0 pg/mL for the lithium heparin and K2 EDTA tube types, respectively. Percent recovery for 2-, 4-, 8-, and 10- fold dilutions with NfL Sample Diluent were all within 80-120% of the neat sample concentration.

**Figure 7.**
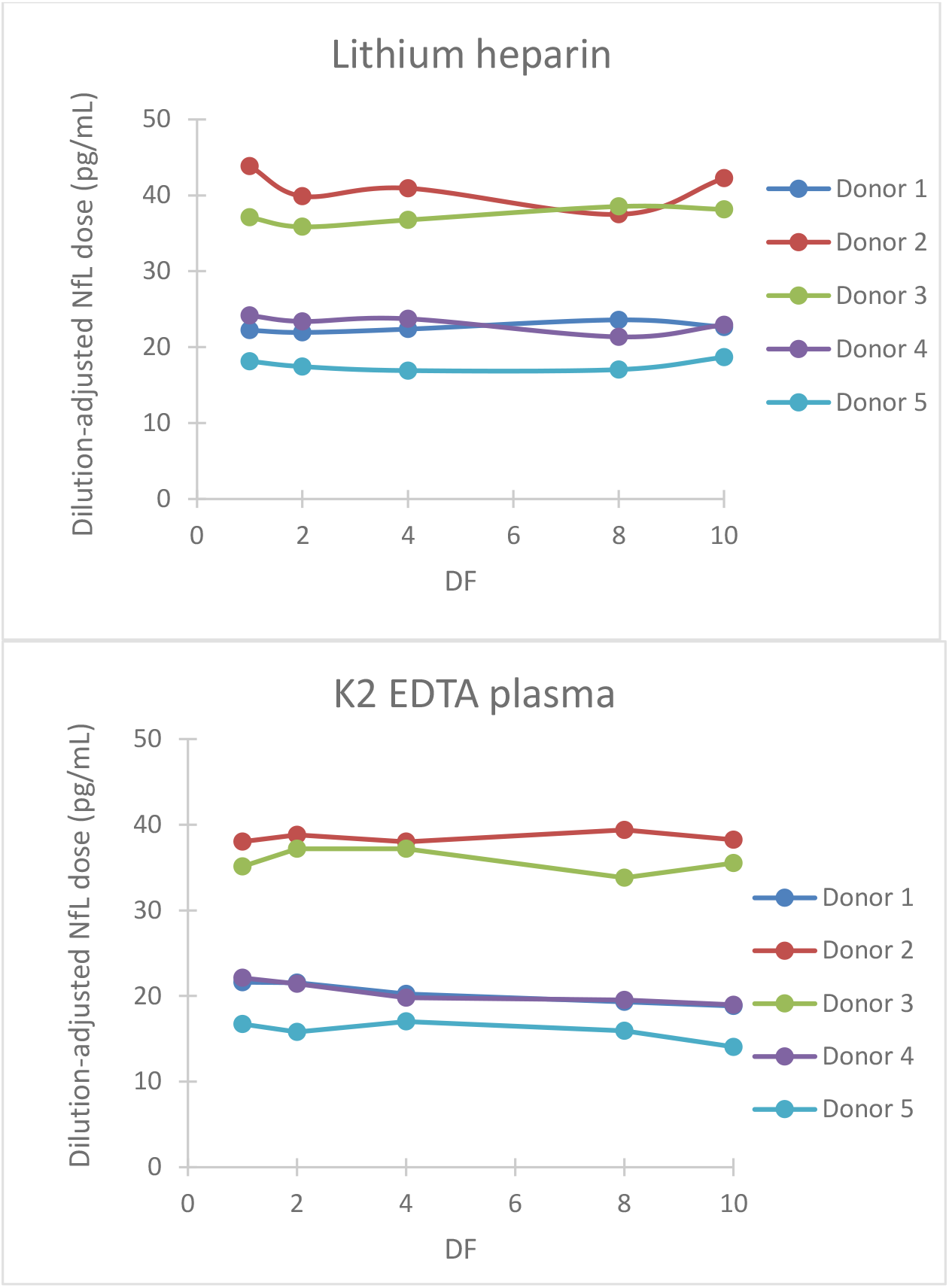
Parallelism for plasma tube types. Five matched lithium heparin and K2 EDTA samples with endogenous NfL levels up to 45 pg/mL were tested neat, and diluted 2-, 4-, 8-, and 10-fold using NfL sample diluent. DF, dilution factor; NfL, neurofilament light chain.

#### CSF Parallelism

Seven CSF samples from normal individuals with NfL levels ranging from 206 - 1439 pg/mL were tested serially diluted 10, 20, 40, 80, 160, and 400-fold using NfL Sample Diluent with the LDT version of the NfL assay. Parallelism was assessed using the 10-fold diluted measured concentration as the expected concentration instead of neat CSF due to 3 of the 4 samples being out of the measurable assay range. All dilutions with measured concentrations above LLoQ for the 7 individual CSF donors tested exhibited 80-120% recovery in comparison to the 10-fold diluted concentration (not shown). Four of the seven CSF samples at starting concentrations less than 400 pg/mL, did not have reportable results at 400-fold dilution.

### Potential Clinical Application

NfL levels in patients with neurodegenerative diseases were shown to correlate with disease severity (9). To examine whether disease-specific trends in NfL using this assay, serum and CSF samples from patients with MS and ALS, respectively, were tested. Serum NfL data from patients with MS demonstrated association with radiological disease activity (11). Baseline sNfL levels from 212 patients with MS were separated into tertiles and compared against the number of new T2 lesions that appeared 6 months later. Analysis of variance demonstrated that patients with a higher NfL level exhibited a statistically significant (*P* < 0.0001) greater number of new T2 lesions after 6 months (**Figure 8A**). NfL levels were also assessed in CSF derived from four healthy controls and four patients with a definite ALS diagnosis. Confirming previous studies (23, 24), NfL was significantly elevated (*P* < 0.0001) in the CSF of patients with ALS (**Figure 8B**). Additionally, NfL levels in CSF were generally two orders of magnitude higher than the levels found in serum (23).

**Figure 8.**
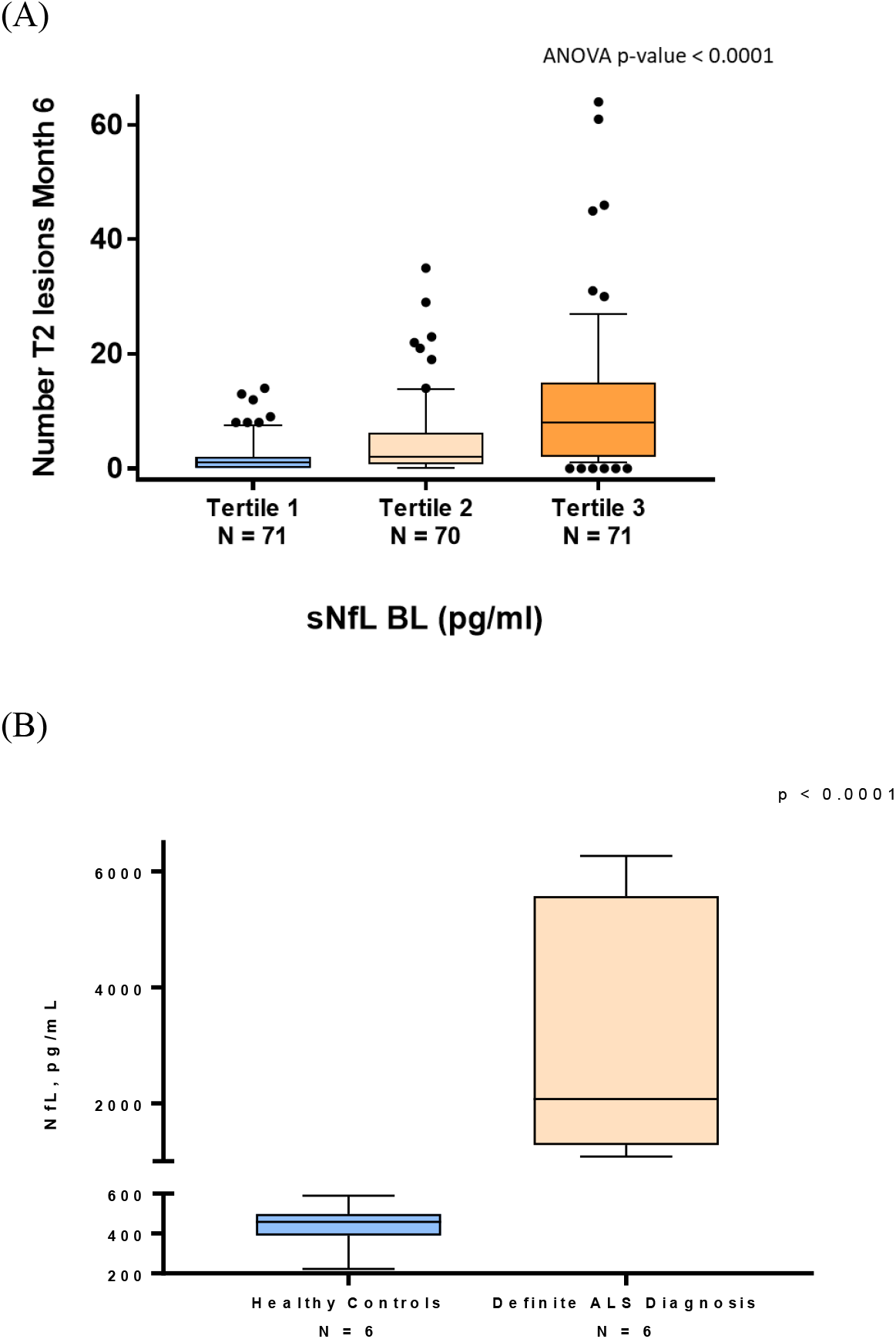
NfL levels are associated with neurodegenerative disease. (A) Patients with MS from the ADVANCE study (11) were separated into tertiles based on baseline (BL) sNfL. The vertical axis shows the number of T2 lesions that developed 6 months later. The lower and upper limit of each tertile were 5.6–11.3 pg/mL for Tertile 1, 11.4–22.1 pg/mL for Tertile 2, and 22.4–100.4 pg/mL for Tertile 3. (B) NfL levels were assessed in CSF derived from healthy controls and patients with a definite ALS diagnosis. ALS, amyotrophic lateral sclerosis; ANOVA, analysis of variance; BL, baseline; CSF, cerebrospinal fluid; NfL, neurofilament light chain; sNfL, serum neurofilament light.

## DISCUSSION

In this report, we describe the performance of a novel research use NfL assay that demonstrates operational and technical features that are compatible with Siemens automated AE-based immunoassay platforms that are used in the laboratory for clinical trials testing applications. The assay provides a wide dynamic range and can be run on plasma, serum, or CSF samples. There is low cross-reactivity with neurofilament medium and heavy chains, and the assay is not significantly affected by various interfering substances encountered in clinical specimens. The NfL assay was designed for compatibility with widely available AE-based platforms. Instrument configurations are available for small, medium, and high-throughput laboratories. In our LDT validation, we utilized the Atellica Solution, which is the highest-throughput platform and the most recently launched hardware; this option would be appropriate for supporting the largest global clinical trials and large clinical practices. Using this platform, the time to first results is 51 minutes and throughput using a single immunoassay module on the Siemens Atellica solution is 171 samples per hour.

Many neurodegenerative diseases often progress stealthily with a long preclinical stage. It is during this prodromal stage that treatments could be most effective before serious, irreversible clinical symptoms become evident. In addition, MRI of the brain has shown that CNS atrophy occurs continuously in diseases such as MS, even during periods of apparent clinical remission. Therefore, it appears that neurodegeneration may be clinically silent in younger patients who are having ongoing low-level CNS injury but can compensate clinically due to reserve capacity and plasticity. However, compensation eventually fails, and the ongoing process accelerates the time to future disability. Therefore, noninvasive biomarkers are needed that can detect underlying pathologies and monitor disease activity. Such biomarkers could also play a role in drug development by providing both a means to stratify patient populations as well as evidence that new drugs are reaching the appropriate molecular target. Furthermore, a biomarker of immune mediate neuronal injury could inform if existing drugs are optimally effective to guide clinical decision-making regarding the need for escalation of disease-modifying immunotherapies of varying potency, as exist for MS. Currently, there is a lack of standardized, validated biomarkers in neurological diseases. NfL, however, is a very promising candidate, with evidence in the literature supporting the value of sNfL as a sensitive and clinically meaningful blood biomarker to monitor neuronal tissue damage and the effects of therapies in neurodegenerative disease (25-29). As we learn more about the strengths and limitations of NfL as a clinical biomarker, it is recognized that a highly sensitive, precise, and accurate test, accessible in the clinical practice setting, would be needed for widespread adoption of NfL in the management of patients with neurodegenerative diseases.

Continuing use of this assay in clinical trials, biomarker validation studies, and with normative reference populations is expected to help establish the utility of NfL in evidence-based decision-making in MS patient care, and as a potential measure of neurodegeneration, which may accelerate development of treatments that slow disease progression in other diseases such as ALS. Demonstration of the performance of a NfL assay on a routine clinical laboratory platform is an important step toward bringing NfL into clinical practice and drug development for a wide range of potential applications in neurology.

## Data Availability

The raw data supporting the conclusions of this article will be made available by the authors, without undue reservation.

## Ethics Statement

The following Ethics Committees and Institutional Review Boards of the participating institutions in Multiple Sclerosis Partners Advancing Technology and Health Solutions (MS PATHS), funded by Biogen, gave ethical approval for this work.

1. University of Rochester Research Subjects Review Board
2. New York University School of Medicine Institutional Review Board
3. Washington University in St. Louis Institutional Review Board
4. Western Institutional Review Board
5. Cleveland Clinic Institutional Review Board, Cleveland Clinic Institutional Review Board
6. Johns Hopkins Medicine Institutional Review Board
7. Ethik-Kommission der Ärztekammer Westfalen-Lippe und der Westfälischen Wilhelms-Universität Münster
8. Comité Ético de Investigación Clínica del Hospital Universitario Vall d’Hebron
9. Ethik-Kommission des Fachbereichs Humanedizin der Philipps-Universität Marburg
10. Ethikkommission an der Technische Universität Dresden

The following Ethics Committees and Institutional Review Boards of the participating institutions in Pegylated interferon β-1a for relapsing-remitting multiple sclerosis (ADVANCE), funded by Biogen, gave ethical approval for this work.

1. Comité d’Ethique UCL Saint-Luc, Avenue Hippocrate 55.14, Tour Havey, niveau 0, Dr. J.M. Maloteaux (Président)/Prof. M.F. van den Hove (Secrétaire), Brussels 1200, Belgium
2. Regionaal Ziekenhuis Sint-Trudo VZW - Ethisch comité, Diestersteenweg 100, Campus Sint-Jozef, Sint-Truiden, 3800, Belgium
3. Ethics Committee for Multi-centre Trials, 5 Sveta Nedelya Sq, Sofia, 1000, Bulgaria
4. University of Western Ontario Health Sciences Research Ethics Board, 1393 Western Road, Room 4180, London, Ontario, N6G 1G9, Canada
5. Comite d’Ethique-Hopital Saint Luc-Edifice Cooper, 3981 Boulevard St. Laurent, Mezzanine 2-Bureau M-207, Montreal, Quebec, Canada
6. Comite de Evaluacion Etico Cientifico del Servicio de Salud Metropolitano Sur Oriente, Avenida Concha y Toro 3459, Oficina de Investigaciones Medicas, Block Central, Puente Alto, Santiago 8207257, Chile
7. EC of Fundacion del Caribe para la Investigacion Biomedica (Fundacion BIOS), Cra 44 No 72 - 131 Oficina 201, Barranquilla, Colombia
8. Comité de Ética e Investigaciones de la Fundación Clínica Abood Shaio, Diagonal 115A Numero 70C-75, Bogota, Colombia
9. Central Ethics Committee, Ksaver 200A, Zagreb, 10000, Croatia
10. Local Drug Committee at Clinical Hospital “Sestre Milosrdnice”, 29 Vinogradska Street, Zagreb, 10000, Croatia
11. Local Drug Committee at Clinical Hospital “Dubrava”, 6 Gojka Suska Avenue, Zagreb, 10000, Croatia
12. Drug Committee Osijek, 4 J.Huttler Street, Osijek, 31000, Croatia
13. Eticka komise Fakultni nemocnice Ostrava, 17. listopadu 1790, Ostrava 708 52, Czech Republic
14. Eticka komise Krajske zdravotni, a.s. - Nemocnice Teplice, o.z., Duchcovska 53, Teplice 415 29, Czech Republic
15. Eticka komise Fakultni nemocnice Olomouc a LF UP v Olomouci, I. P. Pavlova 6, Olomouc 775 20, Czech Republic
16. Eticka komise Fakultni nemocnice v Motole, V Uvalu 84, Praha 5, 150 06, Czech Republic
17. Eticka komise Fakultni nemocnice Brno, Jihlavska 20, Brno 625 00, Czech Republic
18. Eticka komise Vseobecne fakultni nemocnice v Praze, Na Bojisti 1, Praha 2, 128 08, Czech Republic
19. Eticka komise Fakultni nemocnice Brno, Jihlavska 20, Brno 625 00, Czech Republic
20. Tallinn Medical Research Ethics Committee, Hiiu 42, National Institute for Health Development, Room 124, Tallinn EE-11619, Estonia
21. CPP Sud-Méditerranée II - Hôpital Salvator, 249 Boulevard de Sainte Marguerite, Marseille 13009, France
22. Independent Ethics Committee at Ltd “S. Khechinashvili University Clinic, 33, Chavchavadze Ave, Tbilisi 179, Georgia
23. Independent Ethics Committee at Petre Sarajishvili Institute of Neurology, 13, Tevdore Mgvdeli street, Floor 3, Tbilisi 112, Georgia
24. Independent Ethics Committee of Ltd “Research Institute at Clinical Medicine, 13, Tevdore Mgvdeli street, Tbilisi 112, Georgia
25. Independent Ethics Committee at Ltd “Samkurnalo Combinati, 16, Kavtradaze street, Tbilisi 186, Georgia
26. Ethikkommission an der medizinischen Fakultät der Heinrich-Heine-Universität, Moorenstraße 5, Düsseldorf 40225, Germany
27. Ethik-Kommission der Ärztekammer Westfalen-Lippe und der Medizinischen Fakultät der WWU Munster, Von-Esmarch-Straße 62, Medizinische Fakultät der Westfälischen Wilhelms-Universität Münster, Münster 48149, Germany
28. Ethik-Kommission der Bayerischen Landesärztekammer, Mühlbaurstraße 16, München 81677, Germany
29. Ethik-Kommission bei der Ärztekammer Niedersachsen, Berliner Allee 20, Hannover 30175, Germany
30. Landesamt für Gesundheit und Soziales Berlin, Fehrbelliner Platz 1, Geschäftsstelle der Ethikkommission des Landes Berlin, Berlin 10707, Germany
31. Ethikkommission des FB Medizin der Philipps-Universität Marburg, Baldingerstraße, Marburg 35032, Germany
32. Ethik-Kommission an der Medizinischen Fakultät der Universität Leipzig, Härtelstraße 16 – 18, Institut für Klinische Pharmakologie, Leipzig 40107, Germany
33. Ethik-Kommission der Ärztekammer Hamburg, Humboldtstraße 67a, Hamburg 22083, Germany
34. Ethik-Kommission der Medizinischen Fakultät Friedrich-Alexander-Universität Erlangen-Nürnberg, Krankenhausstraße 12, EG, Raum 106, Erlangen 91054, Germany
35. Ethik-Kommission der Medizinischen Hochschule Hannover, Carl-Neuberg-Straße 1, Hannover 30625, Germany
36. Ethik-Kommission der Bayerischen Landesärztekammer, Mühlbaurstraße 16, München 81677, Germany
37. Landesamt für Gesundheit und Soziales Berlin, Fehrbelliner Platz 1, Geschäftsstelle der Ethikkommission des Landes Berlin, Berlin 10707, Germany
38. Ethikkommission der Landesärztekammer Hessen, Im Vogelsgesang 3, Frankfurt 60488, Germany
39. Ethik-Kommission bei der Landesärztekammer Baden-Württemberg, Jahnstraße 40, Stuttgart 70597, Germany
40. Ethikkommission der Medizinischen Fakultät der Ruhr-Universität Bochum, Bürkle-de-la-Camp-Platz 1, BG-Kliniken Bergmannsheil, Bochum 44789, Germany
41. Ethikkommission der Ärztekammer Nordrhein, Tersteegenstraße 9, Düsseldorf 40474, Germany
42. Ethik-Kommission der Medizinischen Fakultät der Ludwig-Maximilians Universität, München, Pettenkoferstr. 8a, München 81675, Germany
43. National Ethics Committee for Clinical Trials, Mesogeion 284, Athens 15562, Greece
44. Institutional Ethics Committee - Deenanath Mangeshkar Hospital, Karve Road, Deenanath Mangeshkar Hospital and Research Centre, 30C, Erandwane, Maharashtra, Pune 411004, India
45. Ethics Committee Poona Hospital Research Centre, 27 Sadashiv Peth, Maharashtra, Pune 411030, India
46. All India Institute of Medical Sciences, 29 Aurobindo Marg, Ansari Nagar, New Delhi 110029, India
47. MCRI Professional Ethics Committee for Research on Human Subjects, Opposite Mahamarg Bus Stand, Mumbai Naka, Curie Manavata Center, Suyojit City Centre, Maharashtra, Nashik 422004, India
48. KMCH Ethics Committee, Avanashi Road, Tamil Nadu, Coimbatore 641014, India
49. Ethics Committee, Jaslok Hospital and Research Centre, 15, Dr.G.Deshmukh Marg, Pedder Road, Maharashtra, Mumbai 400026, India
50. Ethics Committee, Rajinder Nagar, Sir Ganga Ram Hospital, New Delhi 110060, India
51. Ethics Committee Vidyasagar Institute of Mental Health and Neuro-Sciences, 1, Institutional Area, New Delhi, Nehru Nagar 110065, India
52. Institutional Ethics Committee Manipal Hospital and Manipal Heart Foundation, Airport Road, Department of Vascular Surgery, 98, Rustom Bagh Road, Karnataka, Bangalore 560017, India
53. Central India Medical Research Ethics Committee, Dr.S.M.Patil’s Hospital, 2nd Floor, Yugadharma Complex, Ramdaspeth, Nagpur 440010, India
54. Independent Ethics Committee, TN Medical College and BYL Nair Ch. Hospital, Dept of Clinical Pharmacology, Old RMO Bldg, Mumbai 400008, India
55. Max Healthcare Ethics Committee, 1, Press Enclave Road, Saket 110017, India
56. Independent Ethics Committee, Cerebrovascular and Vasculities Research Foundation, Flat ‘B’ Balaji Villa, 9/2, Rajarathinam Street, Kalipauk 600 010, India
57. G.K. Hospital Ethics Committee for Human Subject Research, 11/2 Old Palasia, Department of Neurology, Indore 452018, India
58. Central Ethical Committee, Medical Sciences Complex, Nitte University, Mangalore 575018, India
59. Ethics committee - SMS Medical College and attached Hospital, Jawaharlal Nehru Marg, Jaipur 302004, India
60. Well Care Research Ethics Committee, 25 New Jagnath Road, Behind A.G Office, Gujarat, Rajkot 360001, India
61. Sujlam Independent Ethics Committee, 2nd, floor, AMA House, Near Natraj Cinema, Ashram Road, Ahmedabad 380009, India
62. The Ethics Committee Of Sri Aurobindo Seva Kendra, Sri Aurobindo Seva Kendra, 1H, Gariahat road (South), Kolkata 700068, India
63. SAHEB Central Ethics Committee, Amritsar, 1st floor, 143-144/7, Near Gurunanak Bhawan, City Centre Market, Amritsar 143001, India
64. Bangalore Central Ethics Committee, No. 1423, Kullappa Circle, Kullappa Layout, St. Thomas Town, Kammanahalli, Bangalore 560084, India
65. The Ethics Committee for Clinical Trials on Medicinal Products, Aizkraukles Street 21-113, Riga LV-1006, Latvia
66. Comite de Etica e Investigacion del “Instituto Biomedico de Investigacion A.C.”, Sierra Fria 218, Fraccionamiento Bosques del Prado Norte, Aguascalientes 20217, Mexico
67. Comite de Bioetica del Instituto de Ciencias Biomedicas Angeles, Camino a Santa Teresa 1055, Torre Especialidades, Colonia Heroes de Padierna, Mexico City, DF 10700, Mexico
68. Comite Bioetico para la Investigacion Clinica S.C. Institutional Review Board, Puebla 422 despacho 4, Col. Roma Sur, Mexico, DF 6700, Mexico
69. Comision de Ética del Hospital San José Tec de Monterrey y de la División de Ciencias de la Salud, Av. I. Morones Prieto 3000 Pte, Despacho 1, Col. Los Doctores, Monterrey, Nuevo Leon 64170, Mexico
70. Tijuana General Hospital, Comite de Ensenanza e Investigacion, Aveinda Centenario Numero 10851 Zona Rio, Instituto de Servicios de Salud Publica Del Estado de Baja California, Tijuana, Baja California 22320, Mexico
71. Comité de Ética del Hospital General de Tijuana, Av. Centenario #10851, Zona Rio, C.P. 22320, Tijuana, B.C., Mexico
72. Comité de Ética e Investigación Christus Muguerza del Parque SA de CV, Calle Dr. Pedro Leal Rodriguez 1802, Colonia Centro, Chihuahua 31000, Mexico
73. AZM METC, Oxfordlaan 10, Kamer 4.R1.33, Maastricht 6202 AZ, Netherlands
74. Multi-Region Ethics Committee, PO Box 5013, Level 2, 1-3 The Terrace, Wellington, New Zealand
75. Comite de Etica para la Investigacion de la Universidad de San Martin de Porres Clinica CADAMUJER, Avenida Alameda del Corregidor 1531, Urbanizacion Los Sirius, Las Viñas, La Molina, Lima 12, Peru
76. Comité de Ética en Investigación Biomédica del Hospital Nacional “Dos de Mayo”, Parque Historia de La Medicina Peruana, s/n, Av. Grau, Cuadra 13, Lima, Lima 01, Peru
77. Komisja Bioetyki Uniwersytetu Medycznego w Lodzi, Kosciuszki 4, Lodz 90-419, Poland
78. National Ethics Committee for Clinical Trial on Medicine, 48 Aviator Sanatescu Street, Sector 1, Bucharest 11478, Romania
79. Ethics Committee at the Federal Service on Surveillance in Healthcare and Social Development of RF, Petrovskiy Bulvar, 8, stroenie 2, Moscow 127051, Russia
80. Ethics Committee at Siberian Regional Medical Centre, Ulitsa Kainskaya, 13, Novosibirsk 630007, Russia
81. Ethics Committee within Chelyabinsk City Clinical Hospital #3, Prospect Pobedy, 287, Chelyabinsk 454136, Russia
82. Ethics Committee at Republican Clinical Hospital for Rehabilitation Treatment, Ulitsa Vatutina, 13, Kazan 420021, Russia
83. Ethics Committee within Bashkiria State Medical University, Ulitsa Lenina, 3, Ufa 450000, Russia
84. Ethics Committee within Smolensk Regional Clinical Hospital, Prospect Gagarina, 27, Smolensk 214018, Russia
85. Ethics Committee at City Clinical Hospital # 11, Ulitsa Dvintsev, 6, Moscow 127018, Russia
86. Ethics Committee within Central Clinical Hospital #2 n.a. N.A. Semashko OAO “RZhD”, Ulitsa Budayskaya, 2, Moscow 129128, Russia
87. Ethics Committee within Siberian State Medical University, Moscovskiy Tract, 2, Tomsk 634050, Russia
88. Ethics Committee within Moscow Medical Academy n.a. I.M. Sechenov, 8 Ulitsa Trubetskaya stroenie 2, City Clinical Hospital #61, Moscow 119992, Russia
89. Ethics Committee at City Hospital #2 – Kransodar Multispeciality Treatment and Diagnostics Unit, Ulitsa Krasnykh Partizan, 6, korp. 2, Krasnodar 350012, Russia
90. Ethics Committee at Perm State Medical Academy, Ulitsa Kuybysheva, 39, Department of General Surgery, Perm 614990, Russia
91. Ethics Committee at Perm State Medical Academy, Ulitsa Kuybysheva, 39, Department of General Surgery, Perm 614990, Russia
92. Ethics Committee within Research Institute of Neurology of RAMS, Volokolamskoye Shosse, 80, Moscow 125367, Russia
93. Local Ethics Comittee at Clinical Center NIS, 48 Zorana Djindjica Boulevard, Nis 18000, Serbia
94. Local Ethics Committee at Military Medical Academy, 17 Crnotravska Street, Belgrade 11000, Serbia
95. Local Ethics Comittee at Clinical Center of Serbia, 2 Pasterova Street, Belgrade 11000, Serbia
96. Local Ethics Comittee at Clinical Hospital Center “Kragujevac”, 30 Zmaj Jovina Street, Kragujevac 34000, Serbia
97. CEIC Hospital La Paz, Paseo de la Castellana, 261, Hospital General - Comité Ético de Investigación Clínica, Planta 8, Madrid 28046, Spain
98. CEIC de Andalucía, Avenida de la Innovación s/n, Servicio de Investigación y Desarrollo Personal, Edificio Arena 1, Consejería de Salud - Dirección General de Procesos y Formación, Sevilla 41020, Spain
99. Centro de Farmacovigilancia de Andalucia, Avenida Manuel Siurot, s/n, Servicio de Farmacologia Clinica, Edificio de Laboratorios - 1^a^ Planta, Hospitales Universitarios Virgen del Rocio, Sevilla 41013, Spain
100. CEIC Hospital Virgen Macarena, Calle Dr. Fedriani, 3, Comité de Ensayos Clínicos, Planta 2, Sevilla 41009, Spain
101. CEIC Hospital Universitario Reina Sofía, Avenida Menéndez Pidal, s/n, Planta 1, Edificio de Consultas Externas, Córdoba 14004, Spain
102. Instituto de Investigación Hospital 12 de Octubre (i+12), Avenida de Córdoba s/n, Area de Gestión de Proyectos - Unidad Administrativa CEIC, Planta 6^a^, Centro de Actividades Ambulatorias - Bloque D, Madrid 28041, Spain
103. Central Commission on Ethics Questions of the MoH of Ukraine, Vulytsya Narodnogo Opolchennya, 5, Kyiv 3680, Ukraine
104. Committee on Ethic Questions of Kyiv City Clinical Hospital #4, Vulytsya Solomyanska, 17, Kyiv 3110, Ukraine
105. Commission on Ethics Questions of Municipal Institution of Healthcare Kyiv Regional Clinical Hospital, Vulytsya Baggovutivska, 1, Thoraco-Pulmonary Centre, Kyiv 4107, Ukraine
106. Commission on Biomedical Ethics Questions of Chernivtsi Regional Psychiatric Hospital, Vulytsya Musorgskogo, 2, Chernivtsi 58018, Ukraine
107. Commission on Ethics Questions of Crimean Republican Institution Clinical Hospital n.a N.A. Semashko, Vulytsya Kyivska, 69, Simferopol 96017, Ukraine
108. Bioethics Committee within Dnipropetrovsk State Medical Academy, Dzerzhinskogo vulytsya, 9, Dnipropetrovsk 49044, Ukraine
109. Ethics Commission of Ukrainian State Research Institute of Medical and Social Problems of Disability, Provulok Radyanskyy, 1a, Dnipropetrovsk 49027, Ukraine
110. Committee on Medical Ethics of Central Clinical Hospital of Railways of Ukraine, Provulok Balakireva, 5, Kharkiv 61103, Ukraine
111. Commission on Bioethic Questions of Donetsk National Medical University named after M. Horkyy, Prospekt Illicha, 16, Donetsk 83003, Ukraine
112. Comission on Ethics Questions of Vinnytsa Regional Psychoneurological Hospital n.a. Yuschenko, Vulytsya Pyrogova, 109, Vinnytsya 21005, Ukraine
113. Commission on Ethics Questions of Odesa Regional Clinical Hospital, Vulytsya Zabolotnogo, 26, Odesa 65025, Ukraine
114. Commission on Ethics Questions of Poltava Regional Clinical Hospital n.a. M.V. Sklifosovskyy, Vulytsya Shevchenka, 23, Poltava 36024, Ukraine
115. Local Ethic Commission of Institute of Neurology, Psychiatry and Narcology AMS of Ukraine, Vulytsya Akademika Pavlova, 46, Kharkiv 61068, Ukraine
116. Commission on Ethics Questions of Ternopil Regional Municipal Clinical Psychoneurological Hospital, Vulytsya Troleybusna, 14, Ternopil 46027, Ukraine
117. West of Scotland Research Ethics Committee 1, Dumbarton Road, Western Infirmary, Glasgow G11 6NT, United Kingdom
118. Chesapeake Research Review Incorporated Institutional Review Board, 7063 Columbia Gateway Drive, Suite 110, Columbia, MD 21046, United States
119. Johns Hopkins Medicine Institutional Review Board, 1620 McElderry Street, Reed Hall, Suite B-130, Baltimore, MD 21205-1911, United States
120. Western IRB, 3535 Seventh Avenue Southwest, Olympia, WA 98502-5010
121. Saint Joseph’s Hospital and Medical Center Institutional Review Board, 350 West Thomas Road, Phoenix, AZ 85013, United States
122. Mercy Medical Center Institutional Review Committee, 1111 Sixth Avenue, Des Moines, IA 50314, United States
123. Cleveland Clinic Foundation Regulatory Committee, 9500 Euclid Avenue, Desk HSb 103, Cleveland, OH 44195, United States

The Secondary Use Ethics Committee of Biogen gave ethical approval for the use of the ALS samples for this work.

## Author Contributions

All authors contributed to the article as described below, and all approved the submitted version.

SL: conception or design of the work; data collection; data analysis and interpretation; drafting the article; critical revision of the article

TP: conception or design of the work; data collection; data analysis and interpretation; drafting the article; critical revision of the article

CMS: contributed to study design and data review

KX: contributed to study design and data analysis

XQ: conception or design of the work; data collection; data analysis and interpretation

RAR: contributed to study design and manuscript development

PAC: data analysis and interpretation; critical revision of the article

LS: contributed to assay development via data review and suggested experiments and design of validation exercises

DG: contributed to data generation, analysis, and interpretation

DR: contributed to data review and interpretation

CG: conception or design of the work; data collection; data analysis and interpretation

MM: conception or design of the work; data collection; data analysis and interpretation

AJU: conception or design of the work; data analysis and interpretation; drafting the article; critical revision of the article

## Conflict of Interest

SL: employee of Siemens Healthcare Laboratory, LLC

TP: employee of Biogen Inc. at the time of the study, currently employee of Takeda

CMS: employee of Biogen Inc.

KX: employee of Biogen Inc. at the time of the study

XQ: employee of Siemens Healthcare Laboratory, LLC at the time of the study RAR: employee of Biogen Inc. at the time of the study

PAC: none relevant LS: none relevant

DG: employee of Biogen Inc. DR: employee of Biogen Inc.

CG: employee of Siemens Healthcare Laboratory, LLC at the time of the study

MM: employee of Siemens Healthcare Laboratory, LLC at the time of the study

AJU: employee of Siemens Healthcare Laboratory, LLC; has supervised the work of SL, XQ, and MM; and owns shares of Siemens Healthineers AG stock

