## Supplementary Material for "Development of a Highly Sensitive Serum Neurofilament Light Chain Assay on an Automated Immunoassay Platform"

**Supplementary Figure 1.** Serum sample stability with NfL research assay. Measured NfL concentrations of four individuals (serum) with different handling conditions: (A) room temperature up to 48 hours (B) freeze/thawed up to five cycles. NfL, neurofilament light chain.

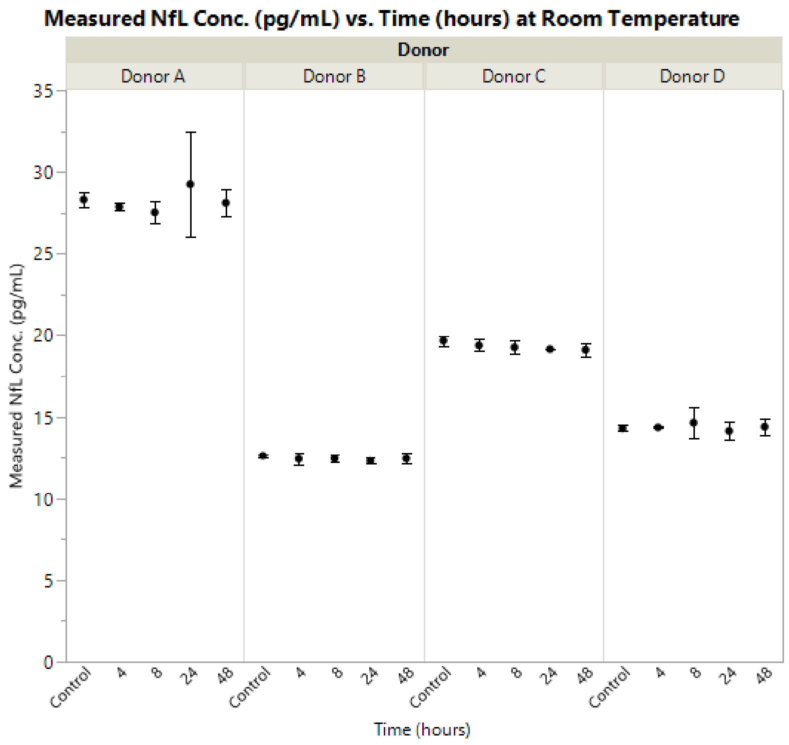


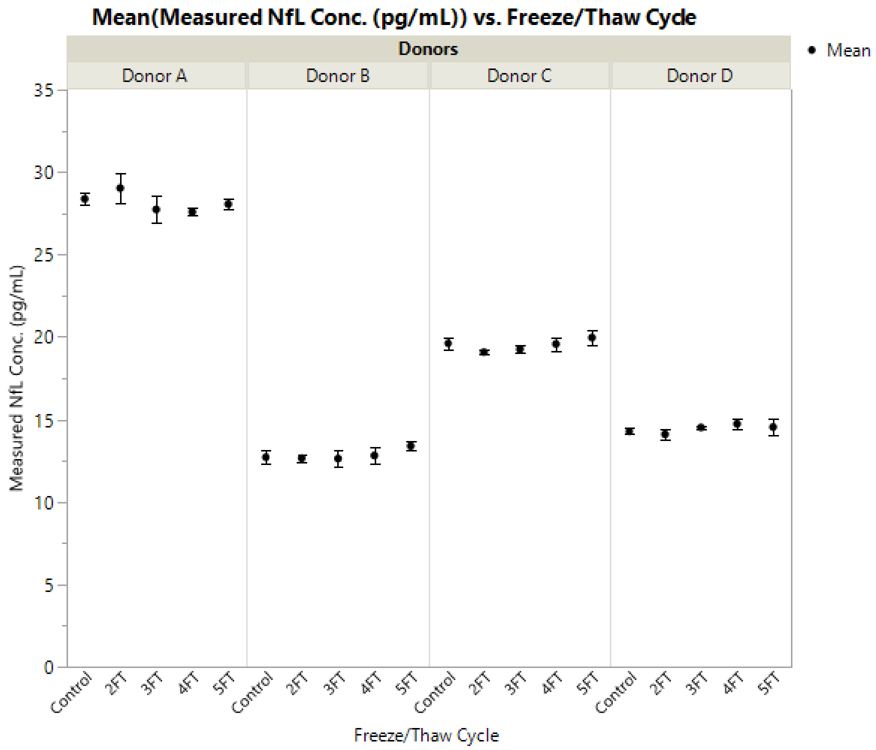

**Supplementary Figure 2.** Hook effect evaluation of NfL research assay. No hook effect observed for NfL assay up to 481 ng/mL (~1000-fold of upper limit of quantification). NfL, neurofilament light chain.

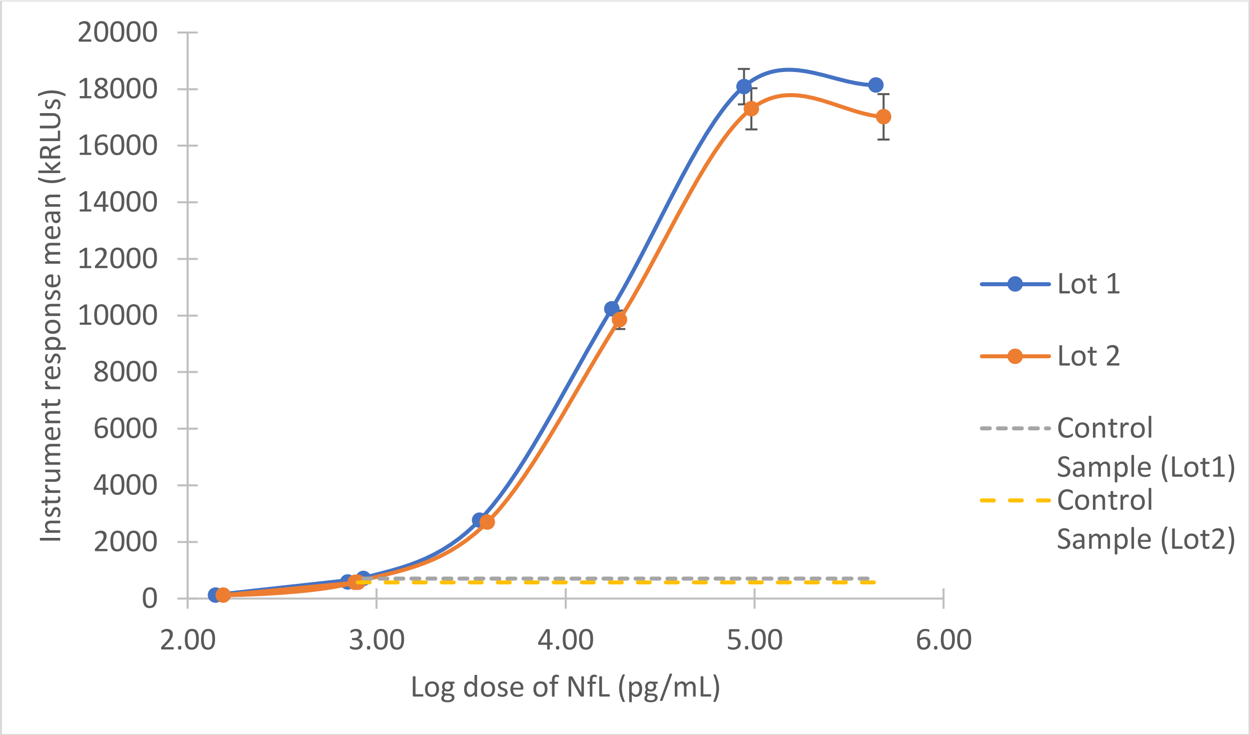
